# Obesity during the COVID-19 pandemic: cause of high risk or an effect of lockdown? A population-based electronic health record analysis in 1 958 184 individuals

**DOI:** 10.1101/2020.06.22.20137182

**Authors:** Michail Katsoulis, Laura Pasea, Alvina G. Lai, Richard JB Dobson, Spiros Denaxas, Harry Hemingway, Amitava Banerjee

## Abstract

**Background:** Obesity is a modifiable risk factor for coronavirus(COVID-19)-related mortality. We estimated excess mortality in obesity, both “direct”, through infection, and “indirect”, through changes in healthcare, and also due to potential increasing obesity during lockdown.

**Methods:** In population-based electronic health records for 1 958 638 individuals in England, we estimated 1-year mortality risk(“direct” and “indirect” effects) for obese individuals, incorporating: (i)pre-COVID-19 risk by age, sex and comorbidities, (ii)population infection rate, and (iii)relative impact on mortality(relative risk, RR: 1.2, 1.5, 2.0 and 3.0). Using causal inference models, we estimated impact of change in body-mass index(BMI) and physical activity during 3-month lockdown on 1-year incidence for high-risk conditions(cardiovascular diseases, CVD; diabetes; chronic obstructive pulmonary disease, COPD and chronic kidney disease, CKD), accounting for confounders.

**Findings:** For severely obese individuals (3.5% at baseline), at 10% population infection rate, we estimated direct impact of 240 and 479 excess deaths in England at RR 1.5 and 2.0 respectively, and indirect effect of 383 to 767 excess deaths, assuming 40% and 80% will be affected at RR=1.2. Due to BMI change during the lockdown, we estimated that 97 755 (5.4%: normal weight to overweight, 5.0%: overweight to obese and 1.3%: obese to severely obese) to 434 104 individuals (15%: normal weight to overweight, 15%: overweight to obese and 6%: obese to severely obese) individuals would be at higher risk for COVID-19 over one year.

**Interpretation:** Prevention of obesity and physical activity are at least as important as physical isolation of severely obese individuals during the pandemic.

Research in context

Evidence before this study
We searched PubMed, medRxiv, bioRxiv, arXiv, and Wellcome Open Research for peer-reviewed articles, preprints, and research reports on obesity, excess mortality and change in body-mass index in the coronavirus disease 2019 (COVID-19), using the search terms “obesity”, “coronavirus”, “COVID-19”, and similar terms, and “mortality”, up to June 15, 2020. We found no prior studies of excess deaths in obese individuals due to COVID-19 pandemic, and no studies of long-term estimates or the relative impact of COVID-19 on mortality. Moreover, there were no studies of change in body-mass index during lockdown periods. Without these data, it is difficult to make specific recommendations in obese people at individual or population level during the pandemic.

Added value of this study
We estimated excess COVID-19-related mortality in severely obese individuals, targeted in physical distancing and isolation policies in UK government guidance. Assuming 10% infection rate, we estimated a direct impact of 240 to 479 excess deaths in England and indirect effect of 383 to 767 excess deaths. On the other hand, we estimated that between 97 755 and 434 104 individuals may develop high-risk conditions for COVID-19 mortality during a 3-month lockdown due to change in body-mass index and physical activity.

Implications of all the available evidence
These analyses support COVID-19 and non-COVID-19 impact assessment in policy planning during the pandemic. The implications of distancing and isolation measures on incidence and mortality from chronic diseases, particularly relating to obesity, needs to be considered in clinical practice, public health and research.

## Introduction

On 13 June 2020, there were 429 878 deaths and 7 800 566 people infected worldwide in the coronavirus (COVID-19) pandemic(1). Over one third of the world’s population (over 3 billion people) is estimated to be under some form of “lockdown”, describing policy-driven, population-level physical isolation to avoid spread of infection(2).

It is clear that the pandemic presents a profound insult *directly* through infection, but also *indirectly* through strain and behaviour change at individual and health system levels(3). Moreover, as countries plan strategies to exit or scale down lockdown, policies to tackle COVID-19 may have unintended consequences on healthcare and outcomes for non-COVID-19 diseases, which we reported for cancer and cardiovascular diseases (CVD)(4,5).

On 16 March 2020, the UK government specified “high-risk” (now “vulnerable”) subgroups for infection(6), moving to the pandemic’s “delay” phase with stringent “physical-distancing” measures. On 22 March 2020, it announced 12 weeks of ‘shielding’ for 1.5 million “extremely vulnerable” people in England with underlying conditions (including severe COPD), but without data underpinning the list of “extremely vulnerable” or “high-risk” conditions(7). On 23 March, the UK lockdown was announced, which is being eased since 11 May.

Among high-risk individuals, severe obesity (body-mass index, BMI ≥40) is a risk factor modifiable in relevant timescales(8). Obesity is common among COVID-19 patients, associated with increasing severity of the disease(9-11). Thus, physical isolation seems reasonable, but impact of age, sex and underlying conditions needs to be better characterised to better tailor policies if further lockdown periods are necessary. Regardless of COVID-19, obesity is associated with increased risk of several chronic conditions, including CVD, diabetes, chronic obstructive airways disease (COPD) and chronic kidney disease (CKD)(12). Prolonged periods of physical isolation could reduce physical activity and increase obesity(13), leading to more people being at high risk during the pandemic.

Using population-based electronic health records (EHR) in England, we estimated: (i) background mortality in severe obesity by underlying risk factors; (ii) direct and indirect excess deaths in individuals with severe obesity; and (iii) impact of BMI gain and physical activity on the incidence of the most common high-risk diseases for COVID-19, and mortality.

## Methods

### Data sources

We used EHR across primary care(Clinical Practice Research Datalink, CPRD-GOLD), hospitals(Hospital Episodes Statistics, HES), and death registry(Office of National Statistics, ONS) with prospective recording and follow-up; linked by CPRD and NHS Digital using unique healthcare identifiers(14). Over 99% of England’s population is general practice(GP)-registered. CPRD is representative by socio-demography, ethnicity and mortality. Approval was by the Independent Scientific Advisory Committee (18_010R) of the UK Medicines and Healthcare products Regulatory Agency in accordance with the Declaration of Helsinki.

### Study population and electronic health record phenotypes

Eligible individuals were 18-69 years old and registered with a GP between 1 January 1998 and 30 June 2016 with BMI and weight data and at least one year of follow-up. Demographic (age, gender, index of multiple deprivation (IMD quintiles and geographic region) and baseline characteristics were recorded at study entry(1 year following latest GP registration)(15). We excluded those ≥70 years of age to focus on high-risk individuals due to obesity alone.

Weight(kg), height(m) and BMI(kg/m^2^) are recorded during GP registration, health checks and at clinical discretion(16). For those with >1 BMI measurement, we selected an eligible BMI record at random during the study period. We excluded BMI records(**Figure S1**): in pregnancy; if 2 same-day observations differed by >0.5 kg/m^2^; if an individual’s highest BMI was more than double their lowest BMI record; and where absolute difference between recorded and calculated BMI on the same date was >1 kg/m^2^. We defined underweight (BMI<18.5kg/m^2^), normal weight (BMI=18.5-24.9kg/m^2^), overweight (BMI=25-29.9kg/m^2^), obesity (BMI≥30kg/m^2^) and severe obesity (BMI≥40kg/m^2)^ and other variables(**Web supplement)**.

### Estimating 1-year mortality

We estimated pre-COVID-19 1-year mortality risk with and without severe obesity using Kaplan-Meier analyses stratified by comorbidities(0, 1, 2 and 3+), scaling from CPRD(N=1,958,184) to the whole English population, aged 18-69 (n=36,621,520 in 2018(17)).

### Estimating 1-year direct and indirect excess deaths in people with severe obesity

Excess deaths were considered as direct(due to or with infection) or indirect(due to changes in health services). *Direct excess deaths* were estimated by applying relative risks, RRs, of 1.2, 1.5, 2 and 3, based on published hazard ratios for CVD and COVID-19 deaths(18, 19), in the absence of cohort studies of clinical cohorts of obese patients investigating all-cause mortality in those with and without infection. We modelled 10% infection rate based on recent seroprevalence estimates(20,21). Although infection rate will change depending on pandemic phase, we assumed infection rate over 1 year in line with the first wave. We used a bias correction factor (incorporating 1-year mortality risk from ONS=0.2912 in 2018) to account for the higher expected mortality in people with at least one BMI measurement in CPRD.

For *direct* and *indirect* excess deaths, we modelled 40% and 80% population affected rates, where 30% and 70% respectively were un-infected but affected at corresponding RRs. Based on RRs from ONS data(1.29 for excess non-COVID-19 deaths) and likely longer-term effects on mortality, we estimated direct and indirect excess deaths together by applying RR 1.2 to 40% and 80% of the population. Thus, we provide *low*(infection rate 10%; no indirect effect at RR 1.5, 2.0 and 3.0), *medium*(infection rate 10%; 30% indirectly affected at RR 1.2) and *high*(infection rate 10%; 70% indirectly affected at RR 1.2) estimates, projected to the whole English population. All analyses were performed using Stata 16MP and R(version 3.4.3).

### Estimating direct and indirect mediators of the effect of severe obesity on mortality

In Table S1, we estimated associations between BMI≥40kg/m^2^ and 1-year mortality using Cox regression models, adjusting for: (1) age and sex; (2) model 1 + 16 combinations of baseline CVD, diabetes, COPD and CKD; and (3) model 2 + baseline hypertension, gout, rheumatoid arthritis and diuretic use. We estimated indirect severe obesity effect by subtracting the model 1 estimate[(Hazard ratio-1)%] from that of models 2 and 3 (i.e. the difference method(22)).

#### Transition to increased BMI group and incidence of high-risk conditions

To calculate 1-year incidence of CVD, COPD, diabetes and CKD, with all and none of the population increasing BMI group, we applied the parametric g-formula, separately in normal weight, overweight and obese individuals(**Figure S2**). Using logistic regression, adjusted for mental health conditions, hypertension, diuretics and cancer (recorded before first BMI measurement and during three months of BMI change), we investigated associations between transition to higher BMI group and 1-year incidence and mortality for the combined outcome. We used coefficients of the model to predict risk of the outcome, if transition was set to zero (i.e. no transition to higher BMI group) and one (i.e. transition to higher BMI group) for all individuals, with standardisation (the mean of all predicted values) and non-parametric bootstrapping from 500 samples to calculate confidence intervals. To estimate incidence of the combined outcome, we assumed different 3-month transition scenarios: normal weight to overweight (5.4% to 15%), overweight to obese (5.0% to 15%) and obese to severely obese(1.3% to 6%)(**Web supplementary methods**). We used 3 months in line with lockdown periods across different countries. In the same logistic regression models, we added physical activity in 4 categories (low, gentle, moderate and vigorous) to estimate its incremental association with 1-year incidence of CVD, diabetes, COPD and CKD.

## Results

### Prevalence of underlying conditions

**Figures S1** and **S2** illustrate the numbers of individuals used in our analyses. The proportion of individuals without comorbidities in individuals without BMI<40kg/m^2^ was 84% and in individuals with BMI≥40kg/m^2^, 70% (**Figure S3**). The prevalence of CVD, diabetes, COPD and CKD and all their combinations were higher among people with severe obesity (**Figure S3**).

### Baseline severe obesity and COVID-19 related excess 1-year mortality

Baseline 1-year mortality risk was 0.83 overall, so we used a bias correction factor of 0.83/0.2912=2.85 in our analysis for the excess deaths calculations (i.e. 0.2912 is the mortality from ONS in 2018 for individuals aged 18-69 years). Mortality at baseline was highest in severely obese individuals age 60-69 with ≥2 underlying conditions(**Figure 1**). The total effect of BMI≥40kg/m^2^ on 1-year mortality is 1.53 (1.42-1.66), after age and sex adjustment (**Table S1**). The proportion of obesity mediated through all combinations of CVD, diabetes, COPD and CKD is 51%. With addition of gout, rheumatoid arthritis, psychiatric diseases and diuretic use, this proportion is 73%.

**Figure 1:**
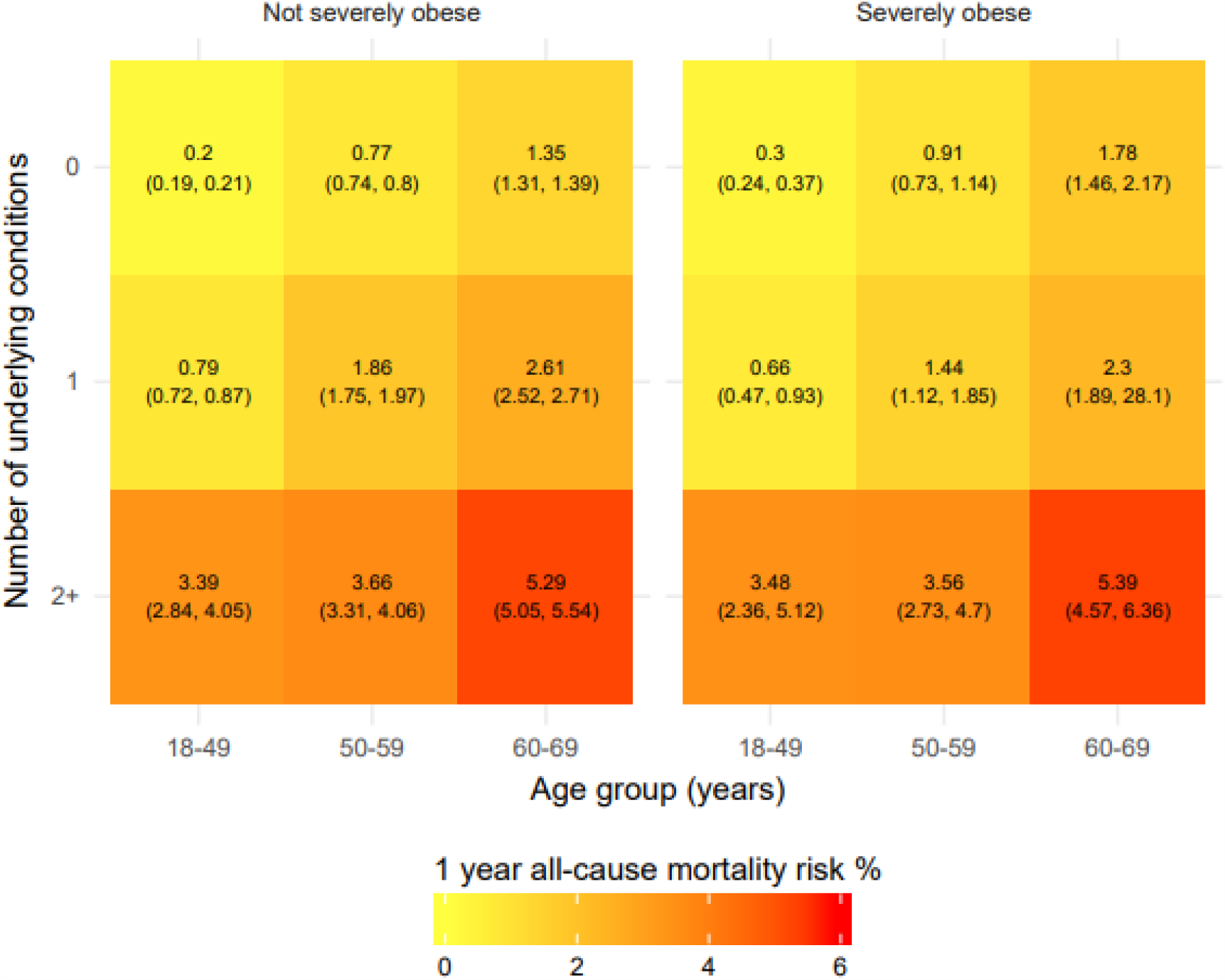
One-year mortality in 1 958 184 individuals in England from the CALIBER study, according to severe obesity, age and number of underlying conditions

At 10% COVID-19 infection rate, there would 240, 479 and 958 excess deaths at RR 1.5, 2.0 and 3.0 respectively in individuals with severe obesity. When 40% and 80% of the population is affected, at RR 1.2, the total excess deaths are estimated to be 383 and 767 respectively for severe obesity**(Figure 2)**.

**Figure 2:**
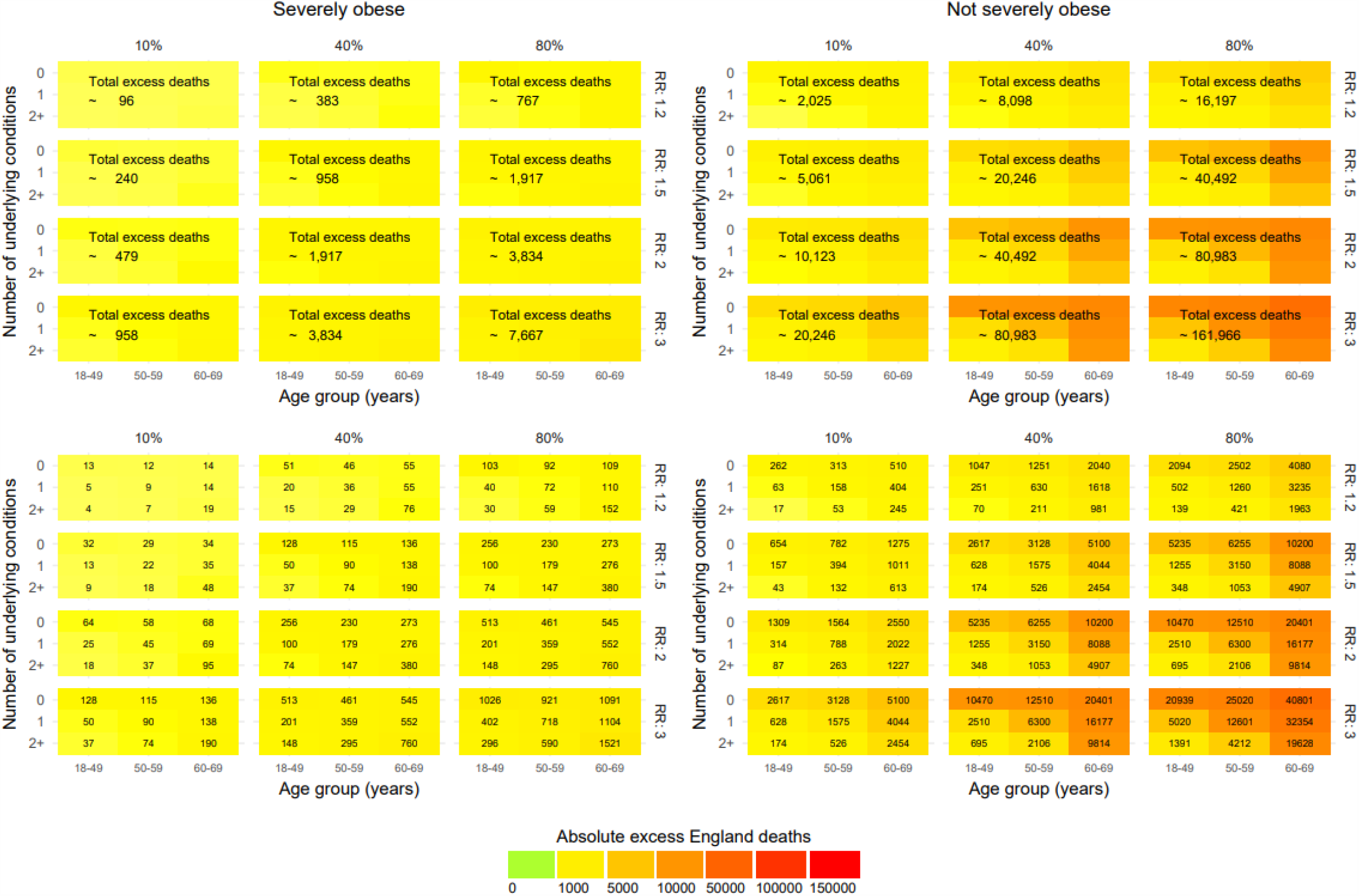
Excess deaths at 1 year in England in different pandemic scenarios by infection rate and relative risks (RR) in people aged 18-69 years old

### BMI transitions and 1-year incidence of high-risk conditions

The risk of transition to a BMI category over 3 months was higher in younger individuals (Figure 3) and for those who were closer to the cutoffs of transition (Figure 3). Transition to a higher BMI group was related to increased risk for incidence of CVD, COPD, diabetes and CKD in 1 year: The odds ratios were 1.07 (0.78-1.48), 1.44 (1.16-1.80) and 1.50 (1.06-2.13) for individuals with normal weight, overweight and obesity respectively **(Table 2**). The risk of developing CVD, diabetes, COPD, and CKD in 1 year, would be 0.68% (0.48-0.90%), 2.35% (1.88-2.80%) and 3.52% (2.39-4.66%) if (due to lockdown) everybody transitioned from normal weight to overweight, from overweight to obesity and from obesity to severe obesity respectively. Conversely, the risk of developing these diseases would be 0.64% (0.58-0.69%), 1.65% (1.56-1.75%) and 2.39% (2.28-2.50%), if normal weight, overweight and obese individuals, respectively, did not transition to higher BMI groups.

**Table 1:** Transition to higher BMI group over 3 months and 1-year incidence of high-risk conditions for COVID-19 ^□^.

|  | **Transition to higher BMI group\* within 3 months vs no transition**<br>Odds ratio (95% CI) | **1-year risk for high-risk conditions for COVID-19<sup>□</sup> (% , 95% CI)** |  |
| --- | --- | --- | --- |
|  |  | **No individuals transition to higher BMI group** | **All individuals transition to higher BMI group** |
| Normal weight<br>N=85 159 | 1.07 (0.78-1.48) | 0.64 (0.58-0.69) | 0.68 (0.48-0.90) |
| Overweight<br>N=81 066 | 1.44 (1.16-1.80) | 1.65 (1.56-1.75) | 2.35 (1.88-2.80) |
| Obese<br>N=87 534 | 1.50 (1.06-2.13) | 2.39 (2.28-2.50) | 3.52 (2.39-4.66) |

\* Transition to higher BMI group: normal weight (BMI: 18.5-24.9kg/m<sup>2</sup>) to overweight (BMI: 25.0-29.9kg/m<sup>2</sup>); overweight (BMI: 25.0-29.9kg/m<sup>2</sup>) to obese (BMI: ≥30kg/m<sup>2</sup>), and obese (BMI: 30.0-39.9kg /m<sup>2</sup>) to severely obese (BMI: ≥40kg/m<sup>2</sup>),
□ Incidence of cardiovascular disease, chronic obstructive pulmonary disease, diabetes and chronic kidney disease in 1 year

**Table 2:** Individuals developing high-risk conditions for COVID-19^*^ in 1-year in England, by BMI group and overall, under different scenarios of transition to higher BMI groups over 3 months

| **BMI group** | **Scenario 1** | **Scenario 2** | **Scenario 3** | **Scenario 4** | **Scenario 5** |
| --- | --- | --- | --- | --- | --- |
|  | Normal weight: 5.3%<br>Overweight: 4.9%<br>Obese: 1.3%<sup>‡</sup> | Normal weight: 8%<br>Overweight: 8%<br>Obese: 2% | Normal weight: 10%<br>Overweight: 10%<br>Obese: 3% | Normal weight: 12%<br>Overweight: 12%<br>Obese: 4% | Normal weight: 15%<br>Overweight: 15%<br>Obese: 6% |
| Normal weight | 325 | 479 | 598 | 718 | 898 |
| Overweight | 4256 | 6865 | 8581 | 10298 | 12872 |
| Obese | 93174 | 140111 | 210167 | 280222 | 420334 |
| Total | 97755 | 147455 | 219346 | 291238 | 434104 |

\* cardiovascular disease, chronic obstructive pulmonary disease, diabetes, chronic kidney disease or severe obesity (BMI≥40kg/m<sup>2</sup>)
<sup>‡</sup>Observed in 2 million individuals in England.

**Figure 3:**
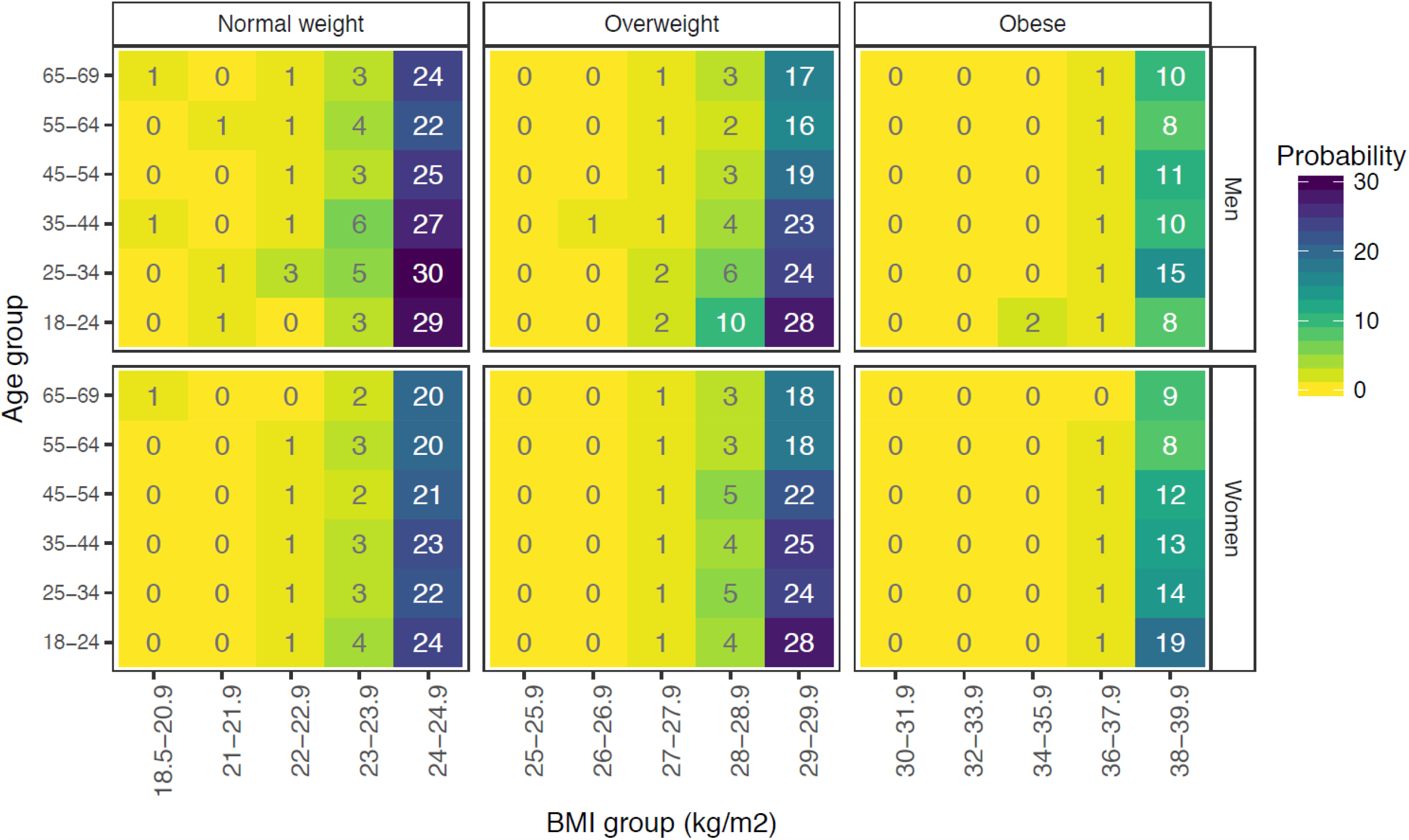
Risk of transition to a higher body mass index (BMI) category over 3 months in 85 159 normal weight, 81 066 overweight and 87 534 obese individuals in England, by age and sex

Assuming that 3-month BMI transitions are in line with average 3-month transitions in CPRD (5.4% from normal weight to overweight, 5% from overweight to obese and 1.3% from obese to severely obese), there would be 97 755 individuals at higher risk for COVID-19 in one year. Assuming higher transition rates of 15% from normal weight to overweight, 15% from overweight to obese and 6% from obese to severely obese respectively, we estimate 434 104 high risk individuals is (**Table 2**).

### Physical activity and 1-year incidence of high-risk conditions

The incidence of high-risk conditions was reduced with moderate physical activity (OR 0.55, 0.30-0.99), but not with gentle (0.71, 0.38-1.31) and vigorous (0.74, 0.35-1.58) activity, in the normal weight group. In overweight individuals, moderate (0.53, 0.38-0.74), and vigorous (0.50, 0.30-0.84) activity were associated with a reduced incidence of high-risk conditions, but not gentle activity (0.76, 0.55-1.06). In obese individuals, gentle (0.78, 0.62-0.97), moderate (0.64, 0.51-0.81), and vigorous (0.49, 0.30-0.81) activity were associated with a reduced incidence of high-risk conditions (**Table 3**). Results remained similar, even after accounting for missing values for physical activity (data not shown).

**Table 3:** Physical activity and 1-year incidence of high-risk conditions for COVID-19 in 85 308 individuals ^*□*^

|  | **Physical activity: Odds ratio (95% CI)** |  |  |  |
| --- | --- | --- | --- | --- |
|  | **Low** | **Gentle** | **Moderate** | **Vigorous** |
| Normal weight<br>N=27,992 | ref | 0.71 (0.38-1.31) | 0.55 (0.30-0.99) | 0.74 (0.35-1.58) |
| Overweight<br>N=28,436 | ref | 0.76 (0.55-1.06) | 0.53 (0.38-0.74) | 0.50 (0.30-0.84) |
| Obese<br>N=28,880 | ref | 0.78 (0.62-0.97) | 0.64 (0.51-0.81) | 0.49 (0.30-0.81) |

<sup>‡</sup> Incidence of cardiovascular disease, chronic obstructive pulmonary disease, diabetes and chronic kidney disease in 1 year

## Discussion

In this large-scale, population-based EHR analysis regarding severe obesity, we investigated excess mortality due to COVID-19 and due to lockdown in England with four main findings. First, we found that severe obesity was generally not associated with high background mortality risk, other than with two or more underlying conditions. Second, relatively few excess deaths are likely to be directly or indirectly associated with severe obesity, compared with obesity overall. Third, lockdown could result in 97 755 to 434 104 individuals transitioning to “high-risk” groups for COVID-19 infection, with significant excess deaths in future. Fourth, physical activity could be protective against adverse outcomes during lockdown.

Prevalence of obesity is increasing over the last decade in all countries, increasing the public health importance of both baseline and excess COVID-19-related risk(8). In obesity and severe obesity, the risk of diabetes and CVD is two and ten times higher than non-obese people, respectively(23), while weight gain is an independent risk factor for both diabetes and CVD(24). We predict that, as at baseline, most of the excess mortality associated with COVID-19 in severely obese individuals will be in people with multimorbidity which should be the focus for prevention, early recognition and aggressive risk factor management. The management of chronic diseases is likely to be indirectly affected during the pandemic and so must be particularly prioritised in the context of obesity.

The largest UK primary care analysis to-date showed that out of 5683 COVID-19 deaths, 257 were in severely obese individuals (hazard ratio, HR 2.27, 1.99-2.58, compared with non-obese individuals), whereas 1631 were in non-severely obese people (HR 1.56, 1.41-1.73)(19). In the UK intensive care audit, 7.7% of patients were severely obese, whereas 31.3% have non-severe obesity(10). There are currently three postulated mechanisms for increased risk observed with obesity: (i) increased susceptibility to infection (e.g. modification of both innate and adaptive immune responses); (ii) increased risk associated with long-term chronic conditions (e.g. CVD, diabetes, COPD and CKD); and (iii) increased COVID-19-specific effects on obese individuals(25). Although we do not present COVID-19 specific data, our data suggest that obesity and its prevention should be viewed more holistically in terms of preventing onset of chronic diseases, rather than emphasising severe obesity. Moreover, these data also support the global picture where high BMI accounts for 4.0 million deaths, with nearly 40% occurring in non-obese individuals, and more than two-thirds due to CVD(8).

The pandemic is having consequences far beyond infection on individuals, populations and health systems, including reductions in bariatric surgery(26). The rise of childhood obesity in Italy(27) and the potential rise in diabetes in India(28) during lockdown have been described, but to our knowledge, this is the first study to model BMI changes across all BMI categories and major chronic diseases together. We show that BMI increases during lockdown could significantly increase the population at high risk from COVID-19, and also lead to indirect future deaths through these incident diseases.

In the UK, as in other European countries, a study using tracking data from wearables and smartphones showed a dramatic increase in homestay duration, a sharp decrease in maximum distance from home and step count, starting from one week prior to lockdown, there was (z-test statistics = 4·1, p-value < 0·001)(29). Physical activity is reduced in obese people during the pandemic(30), and lifestyle modification has been suggested to be an important adjunct to physical distancing and isolation measures(31). We add that exercise reduces incidence of high-risk conditions across the whole BMI range. Exercise should be encouraged during lockdown but also the impact of lockdown on physical activity should be taken into account as policies to ease lockdown are considered and implemented.

### Strengths and limitations

Key strengths of this study include use of population-based EHR, large sample size, utilisation of contemporary and clinically relevant measurements of weight and height, records of relevant comorbidities and causal inference methods (i.e. the g-formula). Normal weight, overweight and obese individuals have different underlying risk for the occurrence of these chronic conditions, hence it is more appropriate to study them separately. Most observational studies cannot focus on this relationship in this resolution, mainly due to their sample size. Several limitations should be noted. Our observational study cannot exclude unmeasured confounding. Since we use EHR, there may be differences between actual and recorded date of incidence of chronic diseases, which would result in bias due to reverse causation. However, this would actually result in underestimation of BMI gain and development of high-risk conditions, except when BMI loss is a result of the disease itself, which would cause potential increased mortality risk. Using EHR, we made assumptions, e.g. linear trend for weight change between 2 body weight measurements recorded <6 months apart, and the last record for physical activity in the last 4 years.

## Conclusions

Our analyses here suggest that despite high prevalence of obesity and underlying conditions, the actual background and excess COVID-19 related mortality due to severe obesity is unlikely to be high compared with other high-risk conditions and is likely to be mediated through other comorbidities. Burden of chronic diseases, even with a 3-month lockdown, may lead to a greater burden of excess deaths, highlighting avoidance of BMI gain and physical activity as public health priorities during the pandemic.

## Data Availability

CPRD data are available by ISAC application and approval.

## Contributorship

Research question: MK, AB.

Funding: AB, SD, HH

Study design and analysis plan: MK, AB, LP, AL.

Preparation of data, including electronic health record phenotyping in the CALIBER open portal; LP, SD

Statistical analysis: MK, LP, AL

Drafting initial and final versions of manuscript: AB, MK

Critical review of early and final versions of manuscript: all authors

## References

1. Worldometer. Coronavirus Worldwide Graphs. 2020. https://www.worldometers.info/coronavirus/worldwide-graphs/ (accessed 29 April 2020)

2. Business Insider. 29 April 2020. https://www.businessinsider.com/countries-on-lockdown-coronavirus-italy-2020-3?r=US&IR=T (accessed 29 April 2020)

3. Banerjee A, Pasea L, Harris S, Gonzalez-Izquierdo A, Torralbo A, Shallcross L, Noursadeghi M, Pillay D, Sebire N, Holmes C, Pagel C, Wong WK, Langenberg C, Williams B, Denaxas S, Hemingway H. Estimating excess 1-year mortality from COVID-19 according to underlying conditions and age in England: a rapid analysis using NHS health records in 3.8 million adults. Lancet (in Press. Cold Spring Harbor Laboratory Press; 2020.

4. Lai AG, Pasea L, Banerjee A, Denaxas S, Katsoulis M, Chang WH, Williams B, Pillay D, Noursadeghi M, Swanton C, Linch D, Hughes D, Forster MD, Johnson P, Turnbull C, DATA-CAN, Cooper M, Jones M, Pritchard-Jones K, Sullivan R, Lawler M, Hall G, Davie C, Hemingway H. Estimating excess mortality in people with cancer and multimorbidity in the COVID-19 emergency. 2020. Preprint 29/4/2020. https://www.researchgate.net/publication/340984562_Estimating_excess_mortality_i n_people_with_cancer_and_multimorbidity_in_the_COVID-19_emergency

5. Banerjee A, Chen S, Pasea L, Lai A, Katsoulis M, Denaxas S, Nafilyan V, Williams B, Wong WK, Bakhai A, Khunti K, Pillay D, Noursadeghi M, Wu H, Pareek N, Bromage D, Mcdonagh T, Byrne J, Teo JT, Shah A, Humberstone B, Tang LV, Shah ASV, Rubboli A, Guo Y, Hu Y, Sudlow CLM, Lip GYH, Hemingway H. Excess deaths in people with cardiovascular diseases during the COVID-19 pandemic. Medrxiv. Preprint. 2020. Online 11/6/2020. https://www.medrxiv.org/content/10.1101/2020.06.10.20127175v1

6. Public Health England. Guidance on social distancing for everyone in the UK and protecting older people and vulnerable adults. 16 March 2020. https://www.gov.uk/government/publications/covid-19-guidance-on-social-distancing-and-for-vulnerable-people/guidance-on-social-distancing-for-everyone-in-the-uk-and-protecting-older-people-and-vulnerable-adults (accessed 29 April 2020)

7. UK Government. https://www.gov.uk/government/news/major-new-measures-to-protect-people-at-highest-risk-from-coronavirus (accessed 29 April 2020)

8. GBD 2015 Obesity Collaborators, Afshin A, Forouzanfar MH, Reitsma MB, Sur P, Estep K, Lee A, Marczak L, Mokdad AH, Moradi-Lakeh M, Naghavi M, Salama JS, Vos T, Abate KH, Abbafati C, Ahmed MB, Al-Aly Z, Alkerwi A, Al-Raddadi R, Amare AT, Amberbir A, Amegah AK, Amini E, Amrock SM, Anjana RM, Ärnlöv J, Asayesh H, Banerjee A, Barac A, Baye E, Bennett DA, Beyene AS, Biadgilign S, Biryukov S, Bjertness E, Boneya DJ, Campos-Nonato I, Carrero JJ, Cecilio P, Cercy K, Ciobanu LG, Cornaby L, Damtew SA, Dandona L, Dandona R, Dharmaratne SD, Duncan BB, Eshrati B, Esteghamati A, Feigin VL, Fernandes JC, Fürst T, Gebrehiwot TT, Gold A, Gona PN, Goto A, Habtewold TD, Hadush KT, Hafezi-Nejad N, Hay SI, Horino M, Islami F, Kamal R, Kasaeian A, Katikireddi SV, Kengne AP, Kesavachandran CN, Khader YS, Khang YH, Khubchandani J, Kim D, Kim YJ, Kinfu Y, Kosen S, Ku T, Defo BK, Kumar GA, Larson HJ, Leinsalu M, Liang X, Lim SS, Liu P, Lopez AD, Lozano R, Majeed A, Malekzadeh R, Malta DC, Mazidi M, McAlinden C, McGarvey ST, Mengistu DT, Mensah GA, Mensink GBM, Mezgebe HB, Mirrakhimov EM, Mueller UO, Noubiap JJ, Obermeyer CM, Ogbo FA, Owolabi MO, Patton GC, Pourmalek F, Qorbani M, Rafay A, Rai RK, Ranabhat CL, Reinig N, Safiri S, Salomon JA, Sanabria JR, Santos IS, Sartorius B, Sawhney M, Schmidhuber J, Schutte AE, Schmidt MI, Sepanlou SG, Shamsizadeh M, Sheikhbahaei S, Shin MJ, Shiri R, Shiue I, Roba HS, Silva DAS, Silverberg JI, Singh JA, Stranges S, Swaminathan S, Tabarés-Seisdedos R, Tadese F, Tedla BA, Tegegne BS, Terkawi AS, Thakur JS, Tonelli M, Topor-Madry R, Tyrovolas S, Ukwaja KN, Uthman OA, Vaezghasemi M, Vasankari T, Vlassov VV, Vollset SE, Weiderpass E, Werdecker A, Wesana J, Westerman R, Yano Y, Yonemoto N, Yonga G, Zaidi Z, Zenebe ZM, Zipkin B, Murray CJL. Health Effects of Overweight and Obesity in 195 Countries over 25 Years. N Engl J Med. 2017 Jul 6;377(1):13–27.

9. Richardson S, Hirsch JS, Narasimhan M, Crawford JM, McGinn T, Davidson KW; and the Northwell COVID-19 Research Consortium, Barnaby DP, Becker LB, Chelico JD, Cohen SL, Cookingham J, Coppa K, Diefenbach MA, Dominello AJ, Duer-Hefele J, Falzon L, Gitlin J, Hajizadeh N, Harvin TG, Hirschwerk DA, Kim EJ, Kozel ZM, Marrast LM, Mogavero JN, Osorio GA, Qiu M, Zanos TP. Presenting Characteristics, Comorbidities, and Outcomes Among 5700 Patients Hospitalized With COVID-19 in the New York City Area. JAMA. 2020 Apr 22. doi: 10.1001/jama.2020.6775.

10. ICNARC report on COVID-19 in critical care. 29 May 2020 https://www.icnarc.org/Our-Audit/Audits/Cmp/Reports (accessed 8 June 2020)

11. Frühbeck G, Baker JL, Busetto L, Dicker D, Goossens GH, Halford JCG, Handjieva-Darlenska T, Hassapidou M, Holm JC, Lehtinen-Jacks S, Mullerova D, O’Malley G, Sagen JV, Rutter H, Salas XR, Woodward E, Yumuk V, Farpour-Lambert NJ. European Association for the Study of Obesity Position Statement on the Global COVID-19 Pandemic. Obes Facts. 2020 Apr 27:1–5. doi: 10.1159/000508082.

12. Nyberg ST, Batty GD, Pentti J, Virtanen M, Alfredsson L, Fransson EI, Goldberg M, Heikkilä K, Jokela M, Knutsson A, Koskenvuo M, Lallukka T, Leineweber C, Lindbohm JV, Madsen IEH, Magnusson Hanson LL, Nordin M, Oksanen T, Pietiläinen O, Rahkonen O, Rugulies R, Shipley MJ, Stenholm S, Suominen S, Theorell T, Vahtera J, Westerholm PJM, Westerlund H, Zins M, Hamer M, Singh-Manoux A, Bell JA, Ferrie JE, Kivimäki M. Obesity and loss of disease-free years owing to major non-communicable diseases: a multicohort study. Lancet Public Health. 2018 Oct;3(10):e490–e497. doi: 10.1016/S2468-2667(18)30139-7. Epub 2018 Sep 1.

13. Lippi G, Henry BM, Bovo C, Sanchis-Gomar F. Health risks and potential remedies during prolonged lockdowns for coronavirus disease 2019 (COVID-19). Diagnosis (Berl). 2020 Apr 7. pii: /j/dx.ahead-of-print/dx-2020-0041/dx-2020-0041.xml. doi: 10.1515/dx-2020-0041. [Epub ahead of print]

14. Denaxas S, Gonzalez-Izquierdo A, Direk K, Fitzpatrick NK, Fatemifar G, Banerjee A, Dobson RJB, Howe LJ, Kuan V, Lumbers RT, Pasea L, Patel RS, Shah AD, Hingorani AD, Sudlow C, Hemingway H. UK phenomics platform for developing and validating electronic health record phenotypes: CALIBER. J Am Med Inform Assoc. 2019;26(12):1545–59.

15. Shah AD, Langenberg C, Rapsomaniki E, Denaxas S, Pujades-Rodriguez M, Gale CP, Deanfield J, Smeeth L, Timmis A, Hemingway H. Type 2 diabetes and incidence of cardiovascular diseases: a cohort study in 1.9 million people. Lancet Diabetes Endocrinol. 2015;3(2):105–13.

16. Nicholson BD, Aveyard P, Bankhead CR, Hamilton W, Hobbs FDR, Lay-Flurrie S. Determinants and extent of weight recording in UK primary care: an analysis of 5 million adults’ electronic health records from 2000 to 2017. BMC Med. 2019 Nov 29;17(1):222. doi: 10.1186/s12916-019-1446-y.

17. Office for National Statistics. Deaths by single year of age tables, UK. https://www.ons.gov.uk/peoplepopulationandcommunity/birthsdeathsandmarriages/deaths/datasets/deathregistrationssummarytablesenglandandwalesdeathsbysingleyearofagetables Date: Jan 17, 2020. Date accessed: April 16, 2020

18. Chen R, Liang W, Jiang M, Guan W, Zhan C, Wang T, Tang C, Sang L, Liu J, Ni Z, Hu Y, Liu L, Shan H, Lei C, Peng Y, Wei L, Liu Y, Hu Y, Peng P, Wang J, Liu J, Chen Z, Li G, Zheng Z, Qiu S, Luo J, Ye C, Zhu S, Liu X, Cheng L, Ye F, Zheng J, Zhang N, Li Y, He J, Li S, Zhong N; Medical Treatment Expert Group for COVID-19. Risk Factors of Fatal Outcome in Hospitalized Subjects With Coronavirus Disease 2019 From a Nationwide Analysis in China. Chest. 2020 Apr 15:S0012-3692(20)30710-8. doi: 10.1016/j.chest.2020.04.010. Online ahead of print.

19. The OpenSAFELY Collaborative; Williamson E. Walker AJ, Bhaskaran K, Bacon S, Bates C, Morton CE, Curtis HJ, Mehrkar A, Evans D, Inglesby P, Cockburn J, McDonald, HI, MacKenna B, Tomlinson L, Douglas IJ, Rentsch CT, Mathur R, Wong A, Grieve R, Harrison D, Forbes H, Schultze A, Croker R, Parry J, Hester F, Harper S, Perera R, Evans S, Smeeth L, Goldacre B. OpenSAFELY: factors associated with COVID-19-related hospital death in the linked electronic health records of 17 million adult NHS patients. Medrxiv. 7 May 2020. https://www.medrxiv.org/content/10.1101/2020.05.06.20092999v1.full.pdf

20. Office of National Statistics. Coronavirus (COVID-19) Infection Survey. https://www.ons.gov.uk/peoplepopulationandcommunity/healthandsocialcare/conditionsanddiseases/datasets/coronaviruscovid19infectionsurveydata (accessed 1 June 2020)

21. Valenti L, Bergna A, Pelusi S, Facciotti F, Lai A, Tarkowski M, Berzuini A, Caprioli F, Santoro L, Baselli G, Ventura CD, Erba E, Bosari S, Galli M, Zehender G, Prati D. SARS-CoV-2 seroprevalence trends in healthy blood donors during the COVID-19 Milan outbreak. May 31 2020. (accessed 1 June 2020) https://www.medrxiv.org/content/10.1101/2020.05.11.20098442v1

22. VanderWeele TJ. A three-way decomposition of a total effect into direct, indirect, and interactive effects. Epidemiology. 2013 Mar;24(2):224–32

23. Kivimäki M, Kuosma E, Ferrie JE, Luukkonen R, Nyberg ST, Alfredsson L, Batty GD, Brunner EJ, Fransson E, Goldberg M, Knutsson A, Koskenvuo M, Nordin M, Oksanen T, Pentti J, Rugulies R, Shipley MJ, Singh-Manoux A, Steptoe A, Suominen SB, Theorell T, Vahtera J, Virtanen M, Westerholm P, Westerlund H, Zins M, Hamer M, Bell JA, Tabak AG, Jokela M. Overweight, obesity, and risk of cardiometabolic multimorbidity: pooled analysis of individual-level data for 120□813 adults from 16 cohort studies from the USA and Europe. Lancet Public Health. 2017 May 19;2(6):e277–e285. Luzi L, Radaelli MG. Influenza and obesity: its odd relationship and the lessons for COVID-19 pandemic. Acta Diabetol. 2020 Apr 5. doi: 10.1007/s00592-020-01522-8. [Epub ahead of print]

24. Katsoulis M, DeStavola B, Diaz-Ordaz K, Gomes M, Lai A, Lagiou P, Wannamethee G, Tsilidis K, Lumbers T, Denaxas S, Banerjee A, Parisinos C, Batterham R, Langenberg C, Hemingway H. Weight change and the incidence of cardiovascular diseases in adults with normal weight, overweight and obesity without chronic diseases; emulating trials using electronic health records Medrxiv 2020. May 27, 2020 https://www.medrxiv.org/content/10.1101/2020.05.14.20102129v2

25. Sattar N, McInnes IB, McMurray JJV. Obesity a Risk Factor for Severe COVID-19 Infection: Multiple Potential Mechanisms. Circulation. 2020 Apr 22.

26. Rubino F, Cohen RV, Mingrone G, le Roux CW, Mechanick JI, Arterburn DE, Vidal J, Alberti G, Amiel SA, Batterham RL, Bornstein S, Chamseddine G, Del Prato S, Dixon JB, Eckel RH, Hopkins D, McGowan BM, Pan A, Patel A, Pattou F, Schauer PR, Zimmet PZ, Cummings DE. Bariatric and metabolic surgery during and after the COVID-19 pandemic: DSS recommendations for management of surgical candidates and postoperative patients and prioritisation of access to surgery.Lancet Diabetes Endocrinol. 2020 May 7:S2213-8587(20)30157-1.

27. Pietrobelli A, Pecoraro L, Ferruzzi A, Heo M, Faith M, Zoller T, Antoniazzi F, Piacentini G, Fearnbach SN, Heymsfield SB. Effects of COVID-19 Lockdown on Lifestyle Behaviors in Children with Obesity Living in Verona, Italy: A Longitudinal Study. Obesity (Silver Spring). 2020 Apr 30.

28. Ghosal S, Sinha B, Majumder M, Misra A. Estimation of effects of nationwide lockdown for containing coronavirus infection on worsening of glycosylated haemoglobin and increase in diabetes-related complications: A simulation model using multivariate regression analysis. Diabetes Metab Syndr. 2020 Apr 10;14(4):319–323. doi: 10.1016/j.dsx.2020.03.014. [Epub ahead of print]

29. Sun S, Folarin AA, Ranjan Y, Rashid Z, Conde P, Stewart C, Cummins N, Matcham F, Costa GD, Leocani L, Sørensen PS, Buron M, Guerrero AI, Zabalza A, Penninx BWJH, Lamers F, Siddi S, Haro JM, Myin-Germeys I, Rintala A, Narayan VA, Comi G, Hotopf M, Dobson RJB, RADAR-CNS consortium. Using smartphones and wearable devices to monitor behavioural changes during COVID-19. 2020. 17 April 2020. https://arxiv.org/ftp/arxiv/papers/2004/2004.14331.pdf

30. Rogers NT, Waterlow N, Brindle HE, Enria L, Eggo RM, Lees S, Roberts CH. Behavioural change towards reduced intensity physical activity is disproportionately prevalent among adults with serious health issues or self-perception of high risk during the UK COVID-19 lockdown. 2020. Medrxiv. May 18, 2020. https://www.medrxiv.org/content/10.1101/2020.05.12.20098921v1.

31. Ho FH, Celis-Morales CA, Gray SR, Katikireddi SV, Niedzwiedz CL, Hastie C, Lyall DM, Ferguson LD, Berry C, Mackay DF, Gill JMR, Pell JP, Sattar N, Welsh P. Modifiable and non-modifiable risk factors for COVID-19: results from UK Biobank. Medrxiv. 2020. May 2, 2020. https://www.medrxiv.org/content/10.1101/2020.04.28.20083295v1.full.pdf

